# Socio-demographic and knowledge-related determinants of vitamin D supplementation in the context of the COVID-19 pandemic: assessment of an educational intervention

**DOI:** 10.1101/2021.04.21.21255553

**Authors:** Katja Žmitek, Maša Hribar, Živa Lavriša, Hristo Hristov, Anita Kušar, Igor Pravst

## Abstract

Vitamin D is a pro-hormone, essential for musculo-skeletal health, normal immune system, and numerous other body functions. Vitamin D deficiency is considered a risk factor in many conditions, and there is growing evidence of its potential role in the severity of COVID-19 outcomes. However, an alarmingly high prevalence of vitamin D deficiency is reported in many regions, and vitamin D supplementation is commonly recommended, particularly during wintertime. To reduce the risk for vitamin D deficiency in the Slovenian population during the COVID-19 pandemic, we conducted mass media intervention with an educational campaign. The objective of this study was to investigate vitamin D supplementation practices in Slovenia before and during the COVID-19 pandemic, and to determine the effects of the educational intervention on supplementation practices. Two data collections were conducted using an online panel with quota sampling for age, sex, and geographical location. A pre-intervention (N=602, April 2020) and post-intervention (N=606) sampling were done during the first and second COVID-19 lockdown, respectively. We also focused on the identification of different factors connected to vitamin D supplementation, with a particular emphasis on vitamin D-related knowledge. Study results showed significant changes in vitamin D supplementation in the population. Penetration of the supplementation increased from 33% in April to 56% in December 2020. The median daily vitamin D intake in supplement users was 25 µg, with about 95% of supplement users taking safe intake levels below 100 µg/daily. Vitamin D-related knowledge (particularly about dietary sources of vitamin D, the health-related impact of vitamin D, and the prevalence of deficiency) was identified as a key independent predictor of vitamin D supplementation. Based on the study findings, we prepared recommendations, which will enable the development of effective awareness campaigns for increasing supplementation of vitamin D.

## 1. Introduction

Vitamin D is a pro-hormone, essential for musculo-skeletal health, normal immune system, and many other body functions (Autier, Boniol, Pizot, & Mullie, 2014; Zittermann, 2003; Zittermann, Pilz, Hoffmann, & Marz, 2016). This micronutrient is also at the frontier of many debates about possible dietary interventions during the COVID-19 pandemic, which has introduced unique threats to the population and has challenged healthcare systems worldwide.

The worst COVID-19 outcomes and higher mortality rates are reported among immunocompromised subjects, including older adults and malnourished people (Barazzoni et al., 2020). Nutritional risks have been identified as particularly relevant, highlighting the need for nutritional interventions (Liu, Zhang, Mao, Wang, & Hu, 2020). Vitamin D deficiency has been recognized as a possible risk of COVID-19 infection and severe disease outcomes (Meltzer et al., 2020; Merzon et al., 2020), therefore vitamin D supplementation is therefore included in recommendations for nutritional support for COVID-19 patients (Anderson, 2020; Caccialanza et al., 2020; Laviano, Koverech, & Zanetti, 2020; Li et al., 2020). While some researchers highlighted possible role of vitamin D in prevention of acute respiratory tract infections (Derbyshire & Calder, 2021; Grant et al., 2020; Adrian R Martineau et al., 2019; A. R. Martineau et al., 2017) and suggested vitamin D supplementation as a possible therapeutic strategy (C. Annweiler et al., 2020; G. Annweiler et al., 2020; Benskin, 2020; Entrenas Castillo et al., 2020; Kaufman, Niles, Kroll, Bi, & Holick, 2020; Rastogi et al., 2020; Shen, Mei, Zhang, & Xu, 2021; Siuka, Pfeifer, & Pinter, 2020), some are highlighting that available results are not yet fully conclusive (Koch, 2021). While it is clear that well-controlled intervention studies are needed in these areas, the high prevalence of vitamin D deficiency in many populations is a rational cause for concerns – with or without COVID-19 pandemic. Some countries therefore updated their Vitamin D supplementation recommendations recently. In the UK for example, revised governmental advice was issued in April 2020 (during the first COVID-19 lockdown), recommending the use of vitamin D supplements for everyone during the autumn and winter months (Koch, 2021). According to additional UK guidance from December 2020, clinically vulnerable people are now offered a free supply of daily vitamin D supplements for 4 months (UK, 2020).

Vitamin D in the body may come both from dietary sources and from biosynthesis in the skin, triggered by sun exposure, more specifically ultraviolet B (UVB) irradiation. The latter represents the main vitamin D source for most of the population, but the efficiency of vitamin D biosynthesis depends on the latitude, season, and several other environmental and personal factors (O’Neill et al., 2016). The usual biomarker for the assessment of vitamin D status is the serum concentration of 25-hydroxy-vitamin D (25(OH)D). Vitamin D deficiency is typically set at serum 25(OH)D concentrations below 50 nmol/l (IOM, 2011), while some researchers suggest even higher optimal target threshold (Holick et al., 2011; McDonnell et al., 2018). Although it has been assumed that sun exposure during summer is sufficient to avoid severe vitamin D deficiency year-round, it is now known that this is not the case in many geographic areas (O’Neill et al., 2016; Spiro & Buttriss, 2014; Zittermann et al., 2016), including Europe (Cashman et al., 2016). Actually, across the northern hemisphere, at latitudes greater than 40°N, the small amount of UVB in sunlight from October to March is insufficient to initiate effective vitamin D synthesis. Therefore, substantial proportions of the European population rely on dietary vitamin D and body stores to maintain a sufficient vitamin D status during the extended winter season, and quite a high prevalence of vitamin D deficiency is reported in many countries worldwide (Calvo, Whiting, & Barton, 2005; Cashman et al., 2016; Pilz et al., 2018; Spiro & Buttriss, 2014). Alarmingly, in our very recent nationally representative study for Slovenia (Hribar et al., 2020), during the extended winter season vitamin D deficiency was found in about 80% of adults (18–74 years), while almost 40% had a severe vitamin D deficiency, with serum 25(OH)D levels below 30 nmol/l.

Natural foods are very limited sources of vitamin D; the most notable sources are oil-rich fish and egg yolks. Consequently, the dietary intake of vitamin D is low in most countries, except in those where oily fish are consumed in high quantities and those with mandatory fortification of foods with vitamin D (Lanham-New & Wilson, 2016). A large European survey, which included several countries, revealed that the mean daily intake of vitamin D is in most cases below 5 μg (200 IU) (Freisling et al., 2010) and such low intakes were confirmed in other studies (Hilger et al., 2014; Spiro & Buttriss, 2014; Wahl et al., 2012). Systemic food fortification has been implemented in many countries in order to increase dietary intake of vitamin D, for example, in the USA, Canada, Australia, and Finland (Pilz et al., 2018). On the contrary, most European countries do not have formal public health fortification or supplementation policies (Spiro & Buttriss, 2014). Slovenia is also an example of a European country that does not have any formal advice or policies regarding the enrichment of food products with this vitamin, and vitamin D supplementation (10 µg/day) is advised routinely only for infants up to 1 year. Our recent study revealed the penetration of vitamin D supplementation in Slovenian adults (18–74 years) reached about 10% (Hribar et al., 2020), but the design of that study did not enable additional analyses, for example, about insights on the seasonal variations in practices of vitamin D supplementation.

In the absence of mandatory food fortification and/or supplementation policies, supplementation practices in the population are on a voluntary basis. Although many factors influence individual behaviors, knowledge is a crucial factor to consider in the development of health promotion programs (Boland, Irwin, & Johnson, 2015). People are exposed to various types of information from different sources and we would expect that personal vitamin D supplementation decisions depend on the knowledge related to this vitamin. Deschasaux et al. reported that, at least in Europe, people are often confused about the sources as well as health effects of vitamin D (2016). Similar observations were also reported in other studies from different countries (Boland et al., 2015; O’Connor, Glatt, White, & Revuelta Iniesta, 2018; Özel, Cantarero-Arevalo, & Jacobsen, 2020; Tariq, Khan, & Basharat, 2020). Physicians and the media were identified as key information providers on this topic, and it was suggested that health professionals should also be better informed about the health effects of vitamin D, and particularly about the vitamin D deficiency risk factors (Deschasaux et al., 2016). Moreover, the public should receive information that reflects the current knowledge on vitamin D health effects and sources. This could contribute to improved vitamin D status in the population.

Vitamin D-related knowledge has not yet been systematically investigated in the Slovenian population, but the high prevalence of vitamin D deficiency and building evidence about its role in COVID-19 have caused increased attention of mass media for this essential micronutrient, which could have affected not only vitamin D-related knowledge, but also supplementation practices. Monthly frequency of articles mentioning vitamin D in Slovenian mass media in the period 2019-2020 is presented in **Figure 1**. There are visible peaks in media coverage of vitamin D during the first COVID-19 lockdown in March, and particularly in the last quartal of 2020. March 2020 peak corresponds with media communication of physician D. Siuka (Siuka, 2020), who proposed 10-steps in the fight against COVID-19 (vitamin D supplementation was mentioned as one of the steps), another peak can be further observed in October 2020, when vitamin D supplementation recommendations for physicians were published on the web site of Slovenian endocrine society (Pfeifer, Siuka, Pravst, & Ihan, 2020), while a major peak occurred in November, after an educational intervention. A press release (NUTRIS, 2020) was sent to major mass media channels, focused on recent results of the national Nutrihealth study (Hribar et al., 2020) about the wide prevalence of vitamin D deficiency in the Slovenian population.

**Figure 1.**
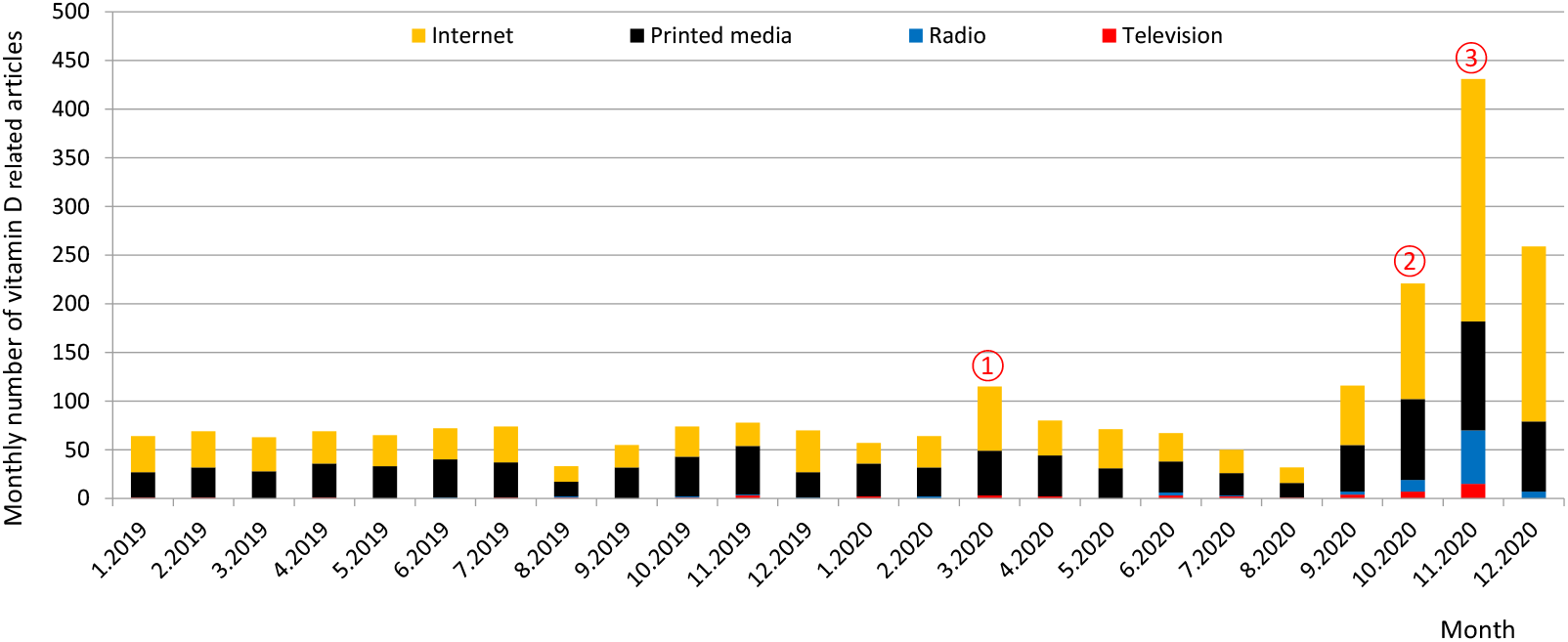
A monthly number of vitamin D-related articles in Slovenian mass media in the years 2019 and 2020. Notes: Figure 1 was constructed using the press coverage data of Kliping media agency (Slovenia), which collects full texts of publications from all relevant mass media channels in Slovenia (covering major television and radio stations (transcripts), print media, and internet portals). Press coverage peaks correspond with: ① Media communication of physician D. Siuka about 10-steps in the fight against COVID-19 (Siuka, 2020); ② Release of recommendations for supplementation with vitamin D for physicians (Pfeifer et al., 2020); ③ Mass media intervention: Press release about the prevalence of vitamin D deficiency in Slovenia by the Nutrition Institute (NUTRIS, 2020), followed by series of interviews and media reports.

Considering these challenges, the objective of this study was to investigate individual vitamin D supplementation practices in Slovenia before and during the COVID-19 pandemic and to determine the effects of the abovementioned educational intervention on supplementation practices. Two data collections were conducted: first during the first COVID-19 lockdown in April 2020, and second during the second COVID-19 lockdown in December 2020 – after an educational intervention. We were focused on the identification of different factors connected to vitamin D supplementation, with a particular emphasis on vitamin D-related personal knowledge. In the absence of mandatory food fortification and vitamin D supplementation, the identification of key knowledge gaps in the population is essential for the preparation of efficient and educational public health campaigns for reducing vitamin D deficiency. Identified knowledge gaps were already used for the educational intervention, which was evaluated with sampling in December 2020.

## 2. Materials and Methods

### 2.1 Data collection

This study was conducted in Slovenia. Sampling was done using an online panel survey in two periods. Frist (pre-intervention) sampling period was between 22nd and 27th April 2020, during the first COVID-19 lockdown, while the second (post-intervention) sampling (N=606) was between 11th and 30th December 2020, during the second COVID-19 lockdown.

Educational intervention (press release to mass media) was done between both collections, on 2nd November 2020 (details provided in Section 2.4). The survey was conducted in the Slovenian language as an amendment to the international Food-COVID-19 survey. Participants provided their informed consent to participate using an online form. Ethical approval for the study was obtained from the Bioethical Committee of the Higher School of Applied Sciences in Ljubljana, Slovenia (VIST ET-6/2020).

Participants were recruited via a consumer panel marketing research institute with quota sampling for age groups, gender, and region. The selected on-line panel (about 35.000 subjects from Slovenia) was used to generate random invitations in the selected quotas. Participants completed the online survey upon invitation. The international Food-COVID-19 questionnaire (Janssen et al., 2021) was amended with additional socio-demographic details (self-reported financial and health status; see details in Section 2.2), with questions about individual vitamin D supplementation practices before and during the COVID-19 pandemic (detail provided in Section 2.2), and vitamin D-related knowledge (detail provided in Section 2.3). Only valid responses for subjects that passed two attention check questions and provided responses to all survey questions are included.

April 2020 pre-intervention sample included N=602 valid responses. Same subjects were also invited to participate in December 2020 post-intervention survey, with a response rate of 62% (n=373). To assure a comparable sample size, additional 233 participants were recruited via the same consumer panel marketing research institute, again with quota sampling for age groups, gender, and region. Complete December 2020 post-intervention sample therefore contained a total of N=606 valid responses.

### 2.2 Variables

Respondents provided information about their age, which, for the purpose of the analysis, was transformed into a categorical variable with four levels: 18–35, 36–49, 50–65, and ≥66 years. Using participants” postal codes and the classification proposed by the European Commission (EUROSTAT, 2020), respondents were classified into three categories: urban, intermediate, and rural. Educational status was also collected using EUROSTAT categorisation (Primary school; Upper secondary – vocational school; Upper secondary – high school; Vocational post-secondary school; First cycle Bologna degree; University or second cycle Bologna degree; Scientific MSc or PhD). For statistical analyses, these categories were joined into three larger education categories—lower (primary school), medium (vocational school or high school), and higher (beyond high school). Self-reported financial status was also measured (“How you would assess financial status of your household”: 1-Very below average; 2-Below average; 3-Average; 4.Above average; 5-Very above average). For statistical analyses, respondents were then classified into three categories: the below average income category (includes respondents indicating very below and below average financial status); the average group (indicates average financial status); and the above average category (indicates above and very above average self-reported financial status). Respondents also reported the size of their household, which was classified into three categories: household with children, single person household, household with 2+ adults without children living together. Self-assessed health condition was surveyed with a question “How you would assess your general health condition (1-very low, 2-low, 3-medium, 4-high, 5-very high). For statistical analyses we created three categories: The first included those with very low- and low health condition, the second included respondents with average health condition and the third included participants indicating high and very high self-reported health condition. Moreover, participants were asked to report if they were supplementing their diet with vitamin D (a) before and (b) during the COVID-19 pandemic. If supplementation was reported, the participant was asked to provide the dosage of vitamin D. As we needed details about the supplementation to enable a calculation of individual daily vitamin D dosage, we used following wording of the question: “Provide the dosage of vitamin D that you used (for example: 1000 IU per day, 100 micrograms per week, 5 drops of Plivit D® per day, etc.). Please provide as much details as possible, to enable us calculation of your daily vitamin D dosage. If possible, check intake of vitamin D on the labelling of the product that you used for supplementation of vitamin D”. Questions were also asked about the extent to which their household had been afflicted with COVID-19 using three questions that asked about infection, isolation or quarantine, and hospitalized members. For the purpose of the analysis, these categories were further joined by one variable with two levels. The participants who responded positively to any of the three questions were classified in the “COVID-19 afflicted households” group, and the remaining respondents in the “COVID-19 not afflicted households” group. The following three questions measured respondents” perceived risk in relation to the disease: (1) the likelihood of any member of your household becoming infected by the virus; (2) the likely severity of the virus for any member of your household; and (3) the level of your anxiety concerning the potential impact of the virus on your household. Participants were asked to score these on a scale from 1 (very low) to 5 (very high).

### 2.3 Vitamin D-related knowledge

Vitamin D-related knowledge was measured using an online tool, developed and described in detail by Boland et al. (Boland et al., 2015). This questionnaire contains questions on the following dimensions of vitamin D: (a) dietary sources; (b) health impact; (c) dietary needs; (d) sun exposure and biosynthesis; (e) other factors of biosynthesis; and (f) prevalence of deficiency. The following modifications of the questionnaire were needed:

1. Translation to Slovenian language.
2. The sun exposure and biosynthesis dimension (d) of the original questionnaire has two questions about the time one needed to spend in the sun to get enough vitamin D—one for fair-skinned persons, and one for non-fair-skinned (i.e., non-Caucasian) persons. We only used the question for fair-skinned persons, as the vast majority of the Slovenian population is Caucasian.
3. The dietary needs (d) of the original questionnaire refer to the daily amount of vitamin D recommended for adults by Health Canada. This question was changed to refer to recommendations applicable in Slovenia, and responses were provided both in International Units and micrograms of vitamin D (only IU in the original questionnaire). While, in the original questionnaire, the correct response was 600 IU (according to Health Canada recommendations), responses of 600 IU/15µg and 800 IU IU/20 µg were considered as correct in our survey, because of the differences between regional and EU-level recommendations (EFSA, 2016; GNS, 2012)

The vitamin D-related knowledge questionnaire used is provided in the Supplementary Information. Six questions were used to assess all six of the above-mentioned dimensions of vitamin D-related knowledge, and each question contributed equally to the calculation of the total knowledge score. Every knowledge question was worth 1 point, producing a maximum score of 6 points. Single questions were scored with 1 if the answer was correct, and 0 if the answer was incorrect. For multiple choice questions, each correct response accounted for a fraction of the overall question. For instance, if a question had 5 correct answers, each contributed 0.2 points. When calculating the total score, the sum of correct responses was deducted from the sum of incorrect responses multiplied by the fraction parts. In this way, a penalty for guessing was implemented to prevent participants from scoring maximum by selecting all possible responses as correct. For this reason, the response “don’t know” was not penalized within the knowledge score. Penalization only occurred within a specific question. In cases where negative scores were given, the whole question was scored as zero.

### 2.4 Educational intervention

Results of pre-intervention data collection in April 2020 were used to identify vitamin D-related knowledge dimensions, connected with vitamin D supplementation practices. Population-based educational intervention started with the launch of a press release on November 2^nd^ 2020, which was sent to e-mail addresses of major Slovenian media channels. The press release was focused on the wide prevalence of vitamin D deficiency in the Slovenian population (NUTRIS, 2020). Intervention resulted in several interviews and numerous publications in mass media. For example, in the last quartal (Q4) of 2020 there were more vitamin D-related publications (N=911) in mass media, than together in the whole year 2019 (N=786) (**Figure 1**).

### 2.5 Data analysis

All statistical analyses were done using STATA version 15.1 (StataCorp LLC, Coledge Station, TX, USA). Descriptive characteristics (mean, median, proportions) are presented for different socio-demographic and individual-based variables and those related to vitamin D supplementation before and during the COVID-19 pandemic. Multivariable linear and logistic regression analyses were used to investigate the predictors of knowledge and supplementation with vitamin D and to determine differences between different sub-populations in terms of knowledge and supplementation. The estimates of vitamin D-related knowledge were determined using age, sex, place of living, education, financial status, health status, and employment, while the estimates of supplementation with vitamin D were determined with respect to age, sex, place of living, education, financial status, and health status. Additionally, a multivariable logistic regression analysis was used to investigate predictors of an increase in vitamin D-related knowledge (December vs. April scoring). In this regard the analyses were conducted using a subsample of subjects, which participated in both pre-intervention (April 2020) and post-intervention (December 2020) data collection, with the exploitation of previously mentioned socio-demographic determinants (age, sex, place of living, education, financial and health status). Multivariable logistic regression analysis was also used to investigate the influence of different dimensions of vitamin D-related knowledge on supplementation with vitamin D, separately for first (April 2020) and second (December 2020) COVID-19 lockdown. For the purpose of binomial regression analysis, respondents were classified into two categories: respondents taking and not taking vitamin D supplements. The model parameters were estimated by the maximum likelihood method. Odds ratios (ORs) with 95% confidence intervals (CIs) were calculated. A z-test for proportions was used to identify significant changes between the pre- and post-intervention supplementation practices. In addition, a t-test for independent samples was used to test the difference in overall vitamin D-related knowledge in pre- and post-intervention sample. Differences were considered significant at p < 0.05.

## 3. Results

### 3.1 Socio-demographic and other characteristics of the sample

Socio-demographic and other characteristics of the participants are presented in Table 1. The compositions of the pre-intervention (April 2020; N=602), the post-intervention (December 2020; N=606), and the combined sample (April and December 2020; N=835) study samples are close to the distribution in the population. Both sex and age distribution are quite comparable, with age groups 19– 35 and 36–49 slightly over-represented, while the 50+ age group is somewhat under-represented. Regarding educational level, the sample is under-represented for the lower education group. Nevertheless, as the study was done as an online survey, such data could not be considered representative, because the population who do not have access to the internet cannot be included.

**Table 1:** Socio-demographic and other characteristics of study participants (Slovenia, 2020).

|  |  | April<br>2020 | December<br>2020 | Combined<br>sample* |
| --- | --- | --- | --- | --- |
| Variables | Levels | N (%) | N (%) | N (%) |
| Sample size |  | 602 (100) | 606 (100) | 835 (100) |
| Age | Mean age in years (SD) | 44.1 (13.5) | 42.94 (13.8) | 41.92 (13.7) |
| Age groups | 18–35 years of age | 179 (29.7) | 206 (34.0) | 301 (36.1) |
|  | 36–49 years of age | 202 (33.6) | 184 (30.4) | 273 (32.7) |
|  | 50–65 years of age | 186 (30.9) | 187 (30.9) | 224 (26.8) |
|  | 66 years and above | 35 (5.8) | 29 (4.8) | 37 (4.4) |
| Sex | Male | 300 (49.8) | 312 (51.5) | 416 (49.8) |
|  | Female | 302 (50.2) | 294 (48.5) | 419 (50.2) |
| Place of living | Urban | 125 (20.8) | 123 (20.3) | 169 (20.2) |
|  | Intermediate | 205 (34.1) | 202 (33.3) | 283 (33.9) |
|  | Rural | 272 (45.2) | 281 (46.4) | 383 (45.9) |
| Education | Primary school | 24 (4.0) | 22 (3.6) | 32 (3.8) |
|  | Upper secondary – vocational school | 77 (12.8) | 52 (8.6) | 92 (11.0) |
|  | Upper secondary – high school | 231 (38.4) | 227 (37.5) | 320 (38.3) |
|  | Vocational post-secondary school | 69 (11.5) | 84 (13.9) | 107 (12.8) |
|  | First cycle Bologna degree | 94 (15.6) | 107 (17.7) | 132 (15.8) |
|  | University or second cycle Bologna degree | 95 (15.8) | 100 (16.5) | 133 (15.9) |
|  | Scientific MSc or PhD | 12 (2.0) | 14 (2.3) | 19 (2.3) |
| Financial status | Very below average | 22 (3.7) | 28 (4.6) | 27 (3.2) |
|  | Below average | 115 (19.1) | 119 (19.6) | 157 (18.8) |
|  | Average | 363 (60.3) | 361 (59.6) | 505 (60.5) |
|  | Above average | 101 (16.8) | 95 (16.7) | 143 (17.1) |
|  | Very above average | 1 (0.2) | 3 (0.5) | 3 (0.4) |
| Health status | Very low | 6 (1.0) | 5 (0.8) | 6 (0.7) |
|  | Low | 17 (2.8) | 18 (3.0) | 21 (2.5) |
|  | Average | 121 (20.1) | 126 (20.8) | 159 (19.0) |
|  | High | 326 (54.2) | 330 (54.5) | 460 (55.1) |
|  | Very high | 132 (21.9) | 127 (21.0) | 189 (22.6) |
| Employment | Full time employed | 318 (52.8) | 331 (54.6) | 449 (53.8) |
|  | Part time employed | 26 (4.3) | 29 (4.8) | 37 (4.4) |
|  | Unemployed | 71 (11.8) | 70 (11.6) | 101 (12.1) |
|  | Keeping house or home maker | 8 (1.3) | 8 (1.3) | 10 (1.2) |
|  | Self-employed | 31 (5.2) | 23 (3.8) | 36 (4.3) |
|  | Student | 49 (8.1) | 58 (9.6) | 92 (11.0) |
|  | Retired | 99 (16.5) | 87 (14.4) | 110 (13.2) |
| Household composition | Household with children | 247 (41.0) | 286 (47.2) | 389 (46.6) |
|  | Single person household | 53 (8.8) | 54 (8.9) | 70 (8.4) |
|  | Household with 2+ adults without children | 302 (50.2) | 266 (43.9) | 376 (45.0) |
| From COVID-19 affected households | Affected | 53 (8.8) | 151 (24.9) | N/A |
|  | Not affected | 549 (91.2) | 455 (75.1) | N/A |
| COVID-19 risk perception:<br>Mean score ± SD | The likelihood of any member of your household becoming infected with the virus. | 2.2 ± 0.9 | 2.7 ± 1.0 | N/A |
| | The likely severity of the virus for any member of your household. | 2.6 ± 1.2 | 2.8 ± 1.2 | N/A |
| | The level of your anxiety concerning the potential impact of the virus on your household. | 2.7 ± 1.1 | 2.9 ± 1.1 | N/A |

**Notes:** SD = standard deviation; N/A = Not applicable COVID-19 risk perception score on scale from 1 (very low) to 5 (very high). \*Combined sample include different participants included in both April and December 2020 samples (373 subjects participated in both April and December surveys, 229 in April study, and 233 in Decembers study only)

The results in Table 1 reveal that about 9% of participants were somehow affected by COVID-19 in April 2020, while the mean COVID-19 risk perception scores were below medium (3). While the proportion of participants from COVID-19-affected households was much higher in December (25%), all mean risk perception scores (rated from 1-very low to 5-very high) were still below scale medium (3). In April the mean score for the likelihood of a household member becoming infected with the virus (2.2 ±0.9) was lower in comparison with the score for the likely severity of the virus for household members, and the score for the level of anxiety concerning the potential impact of the virus on the household (2.6 ±1.2 and 2.7 ±1.1, respectively). This changed in December, when we observed significant increase (p<0.001) in the reported likelihood of a household member becoming infected with the virus in comparison with April (score 2.7 vs. 2.2, respectively). We should also note that almost half of the sample (45.2 and 46.4% in April and December, respectively) was from a rural environment, which might have affected the COVID-19 risk perceptions of study participants.

### 3.2 Vitamin D-related knowledge

The results of the measurements of vitamin D-related knowledge are presented in **Table 2** and **Figure 2**. The maximum vitamin D-related knowledge score would be 6, but in the April 2020 pre-intervention study the highest observed score in our survey was 3.73, with a mean score of only 1.60 (95% CI: 1.53–1.67). The specific dimensions of the vitamin D knowledge provide even more interesting results. Mean scores for dietary vitamin D sources (Q1) and vitamin D’s health impact (Q2) were 0.26 (95% CI: 0.25–0.28), while mean scores for factors affecting the biosynthesis of vitamin D (Q5) were somewhat lower (0.23; 95% CI: 0.21–0.24). The majority of the respondents (54.5%) were aware that the prevalence of vitamin D is above 20%, while the proportion of participants (Q3) knowing their recommended daily intake of vitamin D (15–20 µg) and (Q4) the necessary sun exposure for an average fair-skinned person, when legs and arms are exposed (10–60 minutes per week), was 10% and 28%, respectively.

**Table 2.** Vitamin D-related knowledge score in study samples (N=602; N=606).

| Sampling period | April 2020 | December 2020 |
| --- | --- | --- |
| Variables | Result | Result |
| Number of subjects; N [%] | 602 [100%] | 606 [100%] |
| Knowledge total score: |  |  |
| Mean (95% CI) | 1.60 (1.53–1.67)<sup>a</sup> | 2.21 (2.12–2.90)<sup>b</sup> |
| Vitamin D-related knowledge for all dimensions*: |  |  |
| Food and other sources (Q1)—mean score (95% CI) | 0.26 (0.25–0.28) | 0.28 (0.27–0.30) |
| Health impact (Q2) – mean score (95% CI) | 0.26 (0.25–0.28)<sup>a</sup> | 0.40 (0.38–0.43)<sup>b</sup> |
| Dietary needs (Q3) – N (%) of correct answers | 60.0 (10.0) | 49 (8.2) |
| Sun exposure and biosynthesis (Q4) – N (%) of correct answers | 171 (28.4) | 167 (27.6) |
| Other factors and biosynthesis (Q5) – mean score (95% CI) | 0.23 (0.21–0.24)<sup>a</sup> | 0.45 (0.43–0.47)<sup>b</sup> |
| Deficiency prevalence (Q6) – N (%) of correct answers | 328 (54.5)<sup>a</sup> | 436 (72.0)<sup>b</sup> |

**Notes:** \* Knowledge score consider sum of scores from two types of questions: three multiple choice questions scaled from 0 to 1 and three single choice questions with two discrete options: correct and incorrect. Different letters next to numbers denote significant difference determined with independent sample t-test and z-test for proportions.

**Figure 2.**
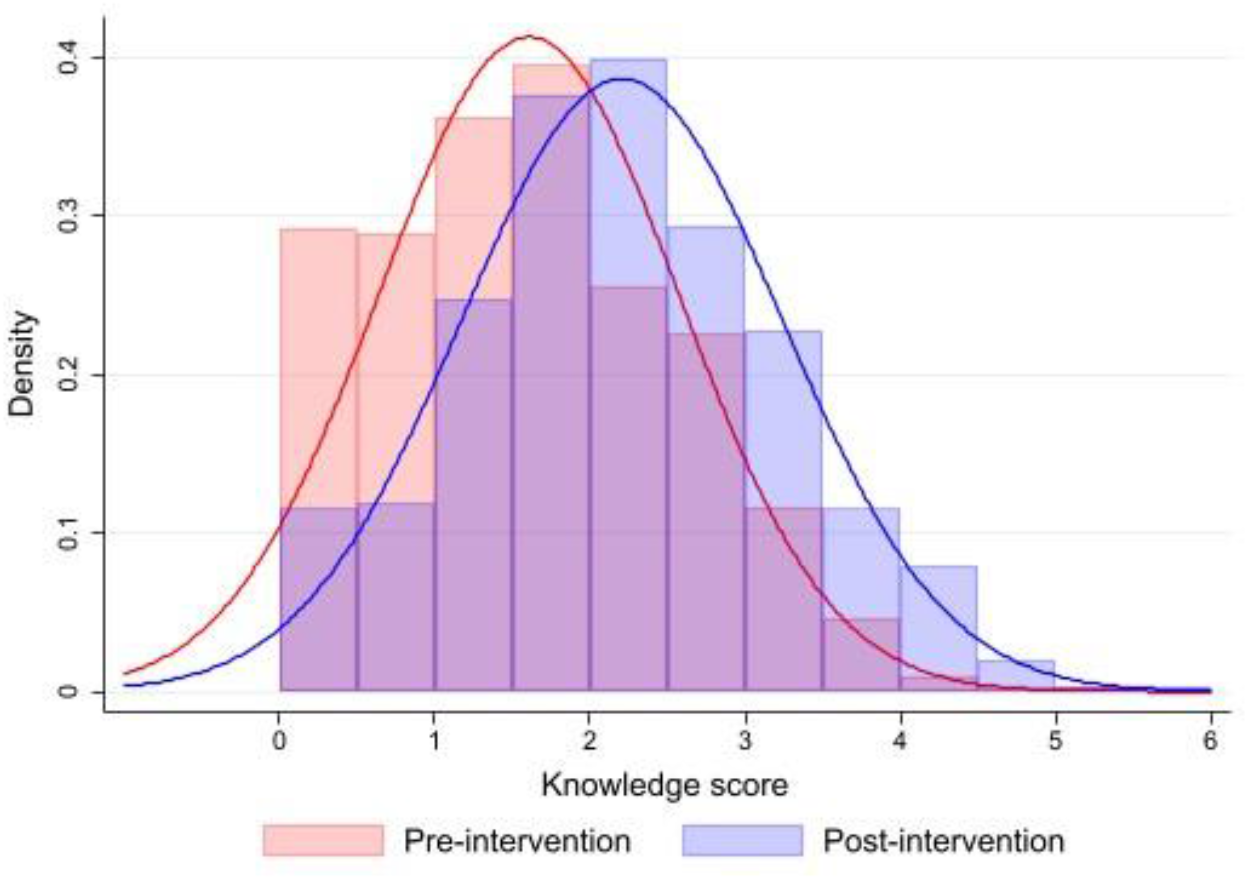
Histograms of pre-intervention (N=602; April 2020) and post-intervention (N=606; December 2020) vitamin D-related knowledge in the study sample. The horizontal (x) scale uses the vitamin D-related knowledge score units, while the respondent’s knowledge distribution is represented by the vertical bars (Red color line depicts normal Gaussian distribution of pre-intervention knowledge score, while blue color line depicts normal Gaussian distribution of post-intervention knowledge score).

December 2020 measurements after conduction of educational intervention showed a statistically significant increase in vitamin-D related knowledge scores (p<0.001). Mean total score significantly increased for 38% and notable differences were also observed in specific dimensions of the vitamin D knowledge (Table 2), particularly in scores for vitamin D’s health impact (Q2: +54%), and factors affecting the biosynthesis of vitamin D (Q5: +96%). As can be observed from knowledge distribution histograms in **Figure 2**, the educational intervention resulted in increase in knowledge on the tail of the distribution, with the population with poor pre-intervention vitamin D-related knowledge having most notable knowledge increase after the intervention.

To provide further insights, linear regression analyses were used to determine adjusted means of vitamin D-related knowledge, considering various socio-demographic and other factors – namely age, sex, place of living, education, financial status, health, and employment status. Analyses were done separately for pre-intervention (April 2020) and post-intervention (December 2020) sample (Table 3). The pre-intervention analysis shows that age, sex and financial status significantly affected vitamin D-related knowledge in different population groups. Older respondents with a higher financial status had significantly increased vitamin D-related knowledge, as shown by the groups’ marginal means. Additionally, significantly higher vitamin D-related knowledge was observed for females compared to the male population. On the other hand, in the post-intervention analyses (December 2020), financial status and sex were not identified as significant factors affecting vitamin D-related knowledge anymore. However, the effect of age was still significant, and the effect of place of living also became significant, with the highest knowledge scores in participants from urban areas.

**Table 3.** Pre-intervention (N=602; April 2020) and post-intervention (N=606; December 2020) adjusted mean (95% CI) levels of vitamin D-related knowledge by age, sex, place of living, education, financial status, health status, and employment.

| Variables | Levels | Pre-intervention (April 2020) |  | Post-intervention (December 2020) |  |
| --- | --- | --- | --- | --- | --- |
|  |  | N (%) | Adjusted | N (%) | Adjusted |
| Overall |  | 602 (100) |  | 606 (100) |  |
| Age groups | 18–35 years of age | 179 (29.7) | 1.43 (1.30–1.58)<sup>a</sup> | 206 (34.0) | 1.99 (1.83–2.15)<sup>a</sup> |
|  | 36–49 years of age | 202 (33.6) | 1.53 (1.39–1.67)<sup>ab</sup> | 184 (30.4) | 2.08 (1.92–2.24)<sup>a</sup> |
|  | 50–65 years of age | 186 (30.9) | 1.80 (1.66–1.95)<sup>b</sup> | 187 (30.9) | 2.52 (2.36–2.68)<sup>b</sup> |
|  | 66 years and above | 35 (5.8) | 1.86 (1.47–2.25)<sup>ab</sup> | 29 (4.8) | 2.53 (2.08–2.97)<sup>ab</sup> |
| Sex | Male | 300 (49.8) | 1.49 (1.39–1.60)<sup>a</sup> | 312 (51.5) | 2.15 (2.03–2.26) |
|  | Female | 302 (50.2) | 1.71 (1.60–1.82)<sup>b</sup> | 294 (48.5) | 2.27 (2.15–2.39) |
| Place of living | Urban | 125 (20.8) | 1.65 (1.48–1.82) | 123 (20.3) | 2.41 (2.23–2.59)<sup>b</sup> |
|  | Intermediate | 205 (34.1) | 1.60 (1.50–1.73) | 202 (33.3) | 2.18 (2.04–2.32)<sup>ab</sup> |
|  | Rural | 272 (45.2) | 1.59 (1.49–1.70) | 281 (46.4) | 2.13 (2.02–2.26)<sup>a</sup> |
| Education | Lower | 24 (4.0) | 1.64 (1.25–2.03) | 22 (3.6) | 1.98 (1.54–2.42) |
|  | Medium | 308 (51.2) | 1.56 (1.46–1.67) | 279 (46.0) | 2.11 (1.99–2.24) |
|  | Higher | 270 (44.9) | 1.65 (1.53–1.77) | 305 (50.3) | 2.31 (2.19–2.42) |
| Financial status | Below average | 137 (22.8) | 1.36 (1.19–1.53)<sup>a</sup> | 147 (24.3) | 2.12 (1.94–2.77) |
|  | Average | 363 (60.3) | 1.65 (1.55–1.75)<sup>b</sup> | 361 (59.6) | 2.22 (2.03–2.41) |
|  | Above average | 102 (16.9) | 1.78 (1.59–1.98)<sup>b</sup> | 98 (16.2) | 2.26 (2.06–2.47) |
| Health status | Low | 23 (3.8) | 2.04 (1.64–2.44) | 23 (3.8) | 2.35 (1.93–2.77) |
|  | Average | 121 (20.1) | 1.65 (1.47–1.83) | 126 (20.8) | 2.22 (2.03–2.41) |
|  | High | 458 (76.1) | 1.57 (1.48–1.66) | 457 (75.4) | 2.19 (2.10–2.29) |
| Employment | Employed | 375 (62.3) | 1.63 (1.53–1.73) | 383 (63.2) | 2.16 (2.05–2.27) |
|  | Unemployed | 79 (13.1) | 1.43 (1.19–1.68) | 87 (14.4) | 2.16 (1.88–2.43) |
|  | Student | 49 (8.1) | 1.70 (1.40–2.01) | 58 (9.6) | 2.58 (2.26–2.88) |
|  | Retired | 99 (16.5) | 1.64 (1.43–1.86) | 78 (12.9) | 2.19 (1.95–2.43) |

**Note:** Identified factors based on contrast of marginal linear predictions accounting for vitamin D-related knowledge: (1) April 2020 sample: $p < 0.01$ (age); $p < 0.01$ (sex), $p < 0.01$ (financial status), $p < 0.1$ (health status); (2) December 2020 sample: $p < 0.01$ (age); $p < 0.05$ (place of living), $p < 0.1$ (education), $p < 0.1$ (employment). Predictor levels not sharing the same subscript are significantly different at $p < 0.05$ using pairwise comparisons of predictive margins with Sidak's adjustment method.

Intervention-related changes were additionally investigated using a sample of subjects, for which we had available two measurements of vitamin D-related knowledge. These were N=373 subjects, who collaborated in both pre-intervention (April 2020) and post-intervention (December 2020) surveys. We investigated predictors of an increase in vitamin D-related knowledge using multivariable logistic regression analysis, focusing on age, sex, place of living, education, financial, and health status (Table 4). Altogether, 74% (N=274) of subjects had increased vitamin D-related knowledge scores in December, in comparison with their April 2020 knowledge scoring. Education of the respondents have shown to be the only significant predictor of increased knowledge score.

**Table 4.** Assessment of intervention-related changes in vitamin D-related knowledge by age, sex, place of living, education, financial status, health status and employment (analyses on N=373 subjects, included in both April and December 2020 sampling).

| Variables | Levels | N (%) | Subjects with increase<sup>1</sup> in vitamin-D related knowledge; N (%) | Odds Ratio (CI) |
| --- | --- | --- | --- | --- |
| Overall |  | 373 (100) | 274 (73.5) |  |
| Age groups | 66 years and above | 27 (7.2) | 21 (77.8) | 1.21 (0.40–3.60) |
|  | 50–65 years of age | 144 (38.6) | 109 (75.7) | 1.10 (0.56–2.18) |
|  | 36–49 years of age | 117 (31.4) | 82 (70.1) | 0.86 (0.44–1.68) |
|  | 18–35 years of age | 85 (22.8) | 62 (72.9) | 1 |
| Sex | Female | 177 (47.4) | 135 (76.3) | 1.36 (0.83–2.23) |
|  | Male | 196 (52.6) | 139 (70.9) | 1 |
| Place of living | Urban | 79 (21.2) | 60 (76.0) | 1 |
|  | Intermediate | 124 (33.2) | 95 (76.6) | 1.08 (0.54–2.16) |
|  | Rural | 170 (45.6) | 119 (70.0) | 0.82 (0.43–1.57) |
| Education | Higher | 178 (47.7) | 128 (71.9) | 5.36 (1.61–17.78)<sup>b</sup> |
|  | Medium | 181 (48.5) | 141 (77.9) | 6.34 (1.93–20.78)<sup>b</sup> |
|  | Lower | 14 (3.8) | 5 (35.7) | 1<sup>a</sup> |
| Financial status | Above average | 60 (16.1) | 40 (66.7) | 0.71 (0.31–1.62) |
|  | Average | 225 (60.3) | 170 (75.6) | 1.08 (0.59–1.99) |
|  | Below average | 88 (23.6) | 64 (72.7) | 1 |
| Health status | Low | 13 (3.5) | 10 (76.9) | 1 |
|  | Average | 81 (21.7) | 55 (67.9) | 0.46 (0.10–2.07) |
|  | High | 279 (74.8) | 209 (74.9) | 0.73 (0.17–3.14) |

**Note:** <sup>1</sup>Increase in December 2020 vitamin D-related knowledge score, in comparison with April 2020 scoring. Three respondents showed no change in vitamin D related knowledge. Identified factors based on contrast of marginal linear predictions accounting for increase in vitamin D-related knowledge: $p < 0.01$ (education). Predictor levels not sharing the same subscript are significantly different at $p < 0.05$ using pairwise comparisons of predictive margins with Sidak's adjustment method. Area under receiver operating characteristic (ROC) curve: 0.62.

### 3.3. Vitamin D-supplementation practices

The penetration of the pre-COVID-19 vitamin D supplementation was 33.7%, and very similar also in the early stage of the COVID-19 pandemic in April 2020 (33.2%). Among those participants who reported the amount of vitamin D supplementation they took before the COVID-19 pandemic or during the pandemic in April, 58% did not report any change in their vitamin D supplementation practice, while 21% reported increased vitamin D dosage, and the same percentage (21%) reported reduced vitamin D dosage. Only few subjects reported using daily doses above 100 µg, exceeding the Tolerable Upper Intake Level (UL) of vitamin D (EFSA, 2012). The mean pre- and -mid pandemic daily vitamin D supplementation, calculated after the exclusion of these subjects, was 31.0 µg and 32.2 µg, respectively. Median intake was 25 µg in both cases. Pre-and post-intervention distribution histograms of daily vitamin D dosages are presented in **Figure 3**. It should be noted that we excluded subjects, which did not report daily vitamin D dosage (N=41 in pre-intervention and N=58 in post-intervention), and that first bar represent subjects taking less than 5 µg daily (including those not taking vitamin D). As expected, the distribution is not normal; subjects were typically supplementing vitamin D with standardized pharmaceutical formulations, where most common vitamin D content is 25 µg (1000 IU) per dosage (capsule, tablet,…)(Žmitek, KruŠič, & Pravst, 2021). More exact daily dosages are however also achievable if liquid formulations (such as oil drops) are used.

**Figure 3.**
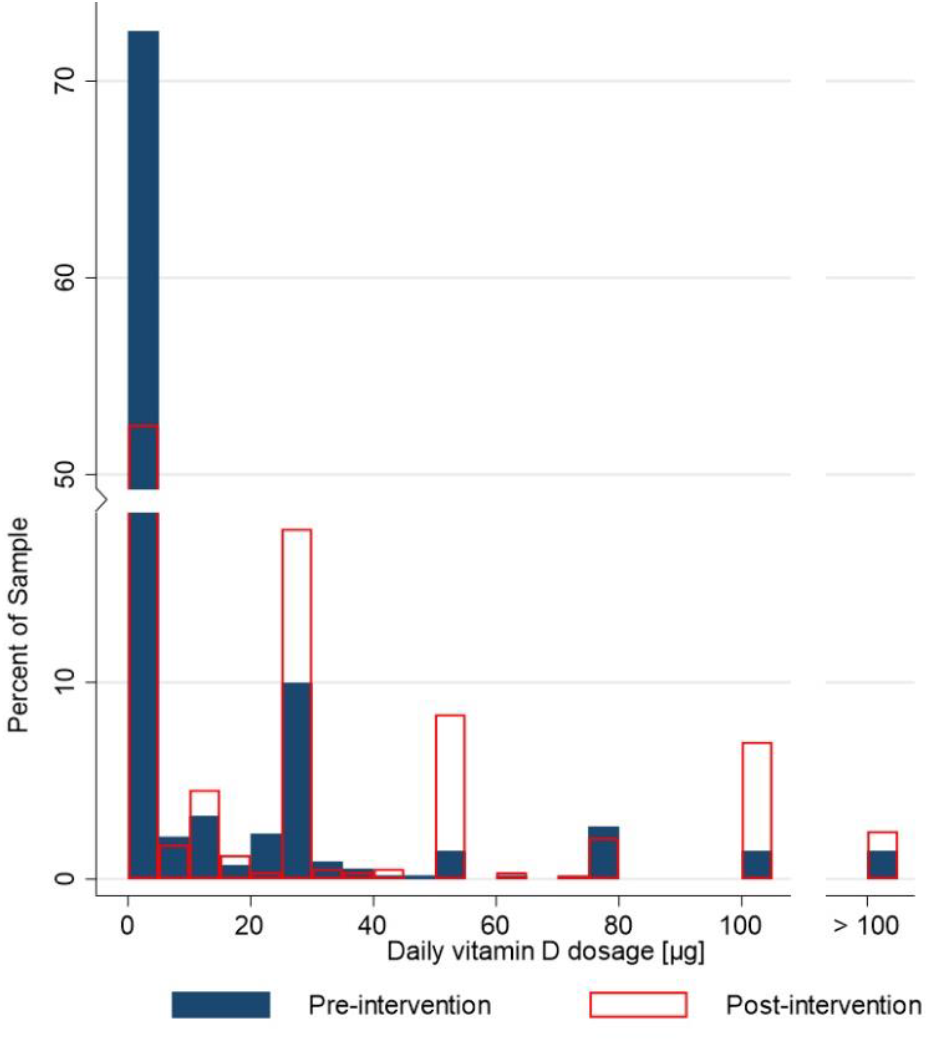
Histogram of pre-intervention (N=573; April 2020: blue) and post-intervention (N=548; December 2020: red) daily vitamin D dosage (µg). The horizontal (x) scale uses daily vitamin D dosage, while the distribution in the population (% of the sample) is represented by the vertical bars. We excluded subjects reporting vitamin D supplementation, which did not report daily vitamin D dosage (N= 41 in pre-intervention and N=58 in post-intervention).

On the other hand, notably different supplementation practices were observed in the later phase of the COVID-19 pandemic, after the educational intervention. Analyses of the post-intervention dataset showed significantly higher penetration of vitamin D supplementation, in comparison to both April and pre-COVID-19 data. Post-intervention proportion of vitamin D supplementation in December 2020 increased considerably for 65%, in comparison to pre-COVID data. While median intake of vitamin D was not changed (25 µg), daily vitamin D supplementation dosage significantly increased to 41.1 µg. Among participants who reported the amount of vitamin D supplementation before the pandemic, or during the December COVID-19 lockdown, 63% reported increased vitamin D dosage, while 20% and 17% reported no change or decrease of daily vitamin D dosage, respectively. The proportion of supplement users exceeding the UL of 100 µg Vitamin D was comparable with April 2020 (5%).

Further we investigated predictors of vitamin D supplementation using multivariable logistic regression analysis, focusing on age, sex, place of living, education, financial and health status (Table 6). Analyses were done separately for pre-intervention (April 2020) and post-intervention (December 2020) samples. In the model, respondents were classified into two categories (respondents supplementing and not supplementing with vitamin D), while model parameters were estimated by the maximum likelihood method. The only two significant predictors were financial and health status. In April 2020, the likelihood for supplementing vitamin D was higher for population with higher financial status and lower health status. Situation changed considerably after the educational intervention, in the December 2020 dataset. Health status was not a significant predictor for vitamin D supplementation anymore, while the financial status was marginally significant (p=0.07). On the other hand, age appear as significant predictor, with the highest vitamin D supplementation rates in elderly subjects.

**Table 5.** Vitamin D supplementation practices before and during the COVID-19 pandemic.

| Variables | Before COVID 19 | During COVID-19<br>Pre-intervention | During COVID-19<br>Post-intervention |
| --- | --- | --- | --- |
| Number of subjects<br>N (%) | 602 (100) | 602 (100) | 606 (100) |
| Reporting Vitamin D supplementation<br>N (%) | 203 (33.7)<sup>a</sup> | 200 (33.2)<sup>a</sup> | 337 (55.6)<sup>b</sup> |
| Reporting daily vitamin D dosage<br>N (%) | 168 (27.9) | 159 (26.4) | 279 (46.0) |
| Daily vitamin D dosage<br>[μg/day] (95% CI)* | 31.0 (27.3–34.7)<sup>a</sup> | 32.2 (28.1–36.2)<sup>a</sup> | 41.1 (37.5–44.7)<sup>b</sup> |
| *Std. Err.* | 1.9 | 2.1 | 1.8 |
| *Median* | 25 | 25 | 25 |
| Subjects with vitamin D<br>dosage above 15 μg/day; N (%) | 125 (74.4) | 124 (95.0) | 231 (82.8) |
| Subjects with vitamin D<br>dosage above 100 μg/day; N(%) | 4 (2.4) | 8 (5.0) | 14 (5.0) |

**Note:** Before COVID-19 pandemic data were collected in April 2020 survey (N=602); \*daily vitamin D dosage was calculated based on the responses of participants who reported the amount of vitamin D supplementation they took before, and during the COVID-19 pandemic (April, December 2020). Different letters next to numbers denote significant difference determined with independent sample t-test and z-test for proportions.

**Table 6.** The proportion of the population using vitamin D supplements during COVID-19 pandemic by age, sex, place of living, education, financial status, and health status: pre-intervention (April 2020) and post-intervention (December 2020) data

| Variables | Levels | *Pre-intervention (April 2020)* |  |  | *Post-intervention (December 2020)* |  |  |
| --- | --- | --- | --- | --- | --- | --- | --- |
|  |  | *N (%)* | Subjects<br>supplementing<br>Vitamin D; *N (%)* | *Odds Ratio (CI)* | *N (%)* | Subjects<br>supplementing<br>Vitamin D; *N (%)* | *Odds Ratio (CI)* |
| Overall |  | 602 (100) | 201 (33.4) |  | 606 (100) | 337 (55.6) |  |
| Age groups | 18–35 years of age | 179 (29.7) | 57 (31.8) | 1 | 206 (34.0) | 101 (49.0) | 1<sup>a</sup> |
|  | 36–49 years of age | 202 (33.6) | 64 (31.7) | 1.00 (0.64–1.56) | 184 (30.4) | 94 (51.1) | 1.13 (0.75–1.70)<sup>ab</sup> |
|  | 50–65 years of age | 186 (30.9) | 66 (35.5) | 1.19 (0.75–1.88) | 187 (30.9) | 121 (64.7) | 1.95 (1.27–3.01)<sup>b</sup> |
|  | 66 years and above | 35 (5.8) | 14 (40.0) | 0.71 (0.74–3.35) | 29 (4.8) | 21 (72.4) | 2.66 (1.09–6.47)<sup>ab</sup> |
| Sex | Male | 300 (49.8) | 95 (31.7) | 1 | 312 (51.5) | 170 (54.5) | 1 |
|  | Female | 302 (50.2) | 106 (35.1) | 1.21 (0.85–1.73) | 294 (48.5) | 167 (56.8) | 1.21 (0.86–1.69) |
| Place of living | Rural | 272 (45.2) | 85 (31.3) | 1 | 281 (46.4) | 152 (54.1) | 1 |
|  | Intermediate | 205 (34.1) | 71 (34.6) | 1.12 (0.75–1.66) | 202 (33.3) | 117 (57.9) | 1.03 (0.71–1.51) |
|  | Urban | 125 (20.8) | 45 (36.0) | 1.19 (0.75–1.89) | 123 (20.3) | 68 (55.3) | 1.00 (0.64–1.54) |
| Education | Lower | 24 (4.00) | 7 (29.2) | 1 | 22 (3.6) | 14 (63.6) | 1 |
|  | Medium | 308 (51.2) | 96 (31.2) | 1.04 (0.41–2.65) | 279 (46.0) | 154 (55.2) | 0.54 (0.21–1.39) |
|  | Higher | 270 (44.6) | 98 (36.3) | 1.22 (0.47–3.14) | 305 (50.3) | 169 (55.4) | 0.55 (0.21–1.43) |
| Financial status | Below average | 137 (22.8) | 34 (24.8) | 1<sup>a</sup> | 147 (24.3) | 77 (52.4) | 1 |
|  | Average | 363 (60.3) | 129 (35.5) | 2.00 (1.24–3.22)<sup>b</sup> | 361 (59.6) | 201 (55.7) | 1.47 (0.96–2.24) |
|  | Above average | 102 (16.9) | 38 (37.3) | 2.20 (1.18–4.09)<sup>b</sup> | 98 (16.2) | 59 (60.2) | 1.91 (1.08–3.41) |
| Health status | High | 458 (76.1) | 146 (31.9) | 1<sup>a</sup> | 457 (75.4) | 241 (52.7) | 1 |
|  | Average | 121 (20.1) | 43 (35.5) | 1.35 (0.86–2.13)<sup>ab</sup> | 126 (20.8) | 80 (63.5) | 1.42 (0.91–2.21) |
|  | Low | 23 (3.8) | 12 (52.2) | 3.05 (1.26–7.40)<sup>b</sup> | 23 (3.8) | 16 (69.6) | 2.16 (0.84–5.53) |

Note: Surveying was done during the first (April 2020) and second (December 2020) COVID-19 lockdown period. Vitamin D-related educational intervention was done between both measurements in November 2020. We identified predictors based on the contrast in marginal linear predictions accounting for vitamin D supplementation: $p = 0.01$ (financial status), $p = 0.03$ (health status) for pre-intervention sample; $p < 0.01$ (age), $p = 0.07$ (financial status) for post-intervention sample. Predictor levels not sharing the same subscript are significantly different using pairwise comparisons of predictive margins with Sidak's adjustment method. Area under receiver operating characteristic (ROC) curve: Pre-intervention: 0.60; post-intervention: 0.62.

To provide further insights into the connection between vitamin D supplementation practices and vitamin D-related knowledge, we used a modeling approach based on the logistic regression method (Figure 4). Two models were constructed to investigate the probability of vitamin D supplementation, using all six investigated dimensions of vitamin D-related knowledge. Model 1 examined pre-intervention vitamin D supplementation practices (April 2020), while Model 2 referred to post-intervention supplementation practices in December 2020. The analysis shows that the increase in vitamin D-related knowledge in three of the six dimensions significantly predicted the likelihood of vitamin D supplementation. The increase in knowledge about dietary sources of vitamin D was found significant predictor in post-intervention Model 2 (OR 2.88, 95% CI: 1.20–6.91, p=0.02), while it was close to significant in pre-intervention model (OR 2.55, 95% CI: 0.98–6.65, p=0.05). On the other hand knowledge about the health-related impact of vitamin D (OR 6.16, 95% CI: 2.89–16.56, p<0.001 in pre-intervention, and OR 4.22, 95%CI: 2.36–7.56, p<0.001 for post-intervention) and knowledge about prevalence of vitamin D deficiency in the population (OR 1.64, 95%CI: 1.10–2.44, p=0.02 before the pandemic, and OR 1.56, 95% CI: 1.06–2.29, p<0.03 during the pandemic) significantly increase the probability of vitamin D supplementation in both models.

**Figure 4.**
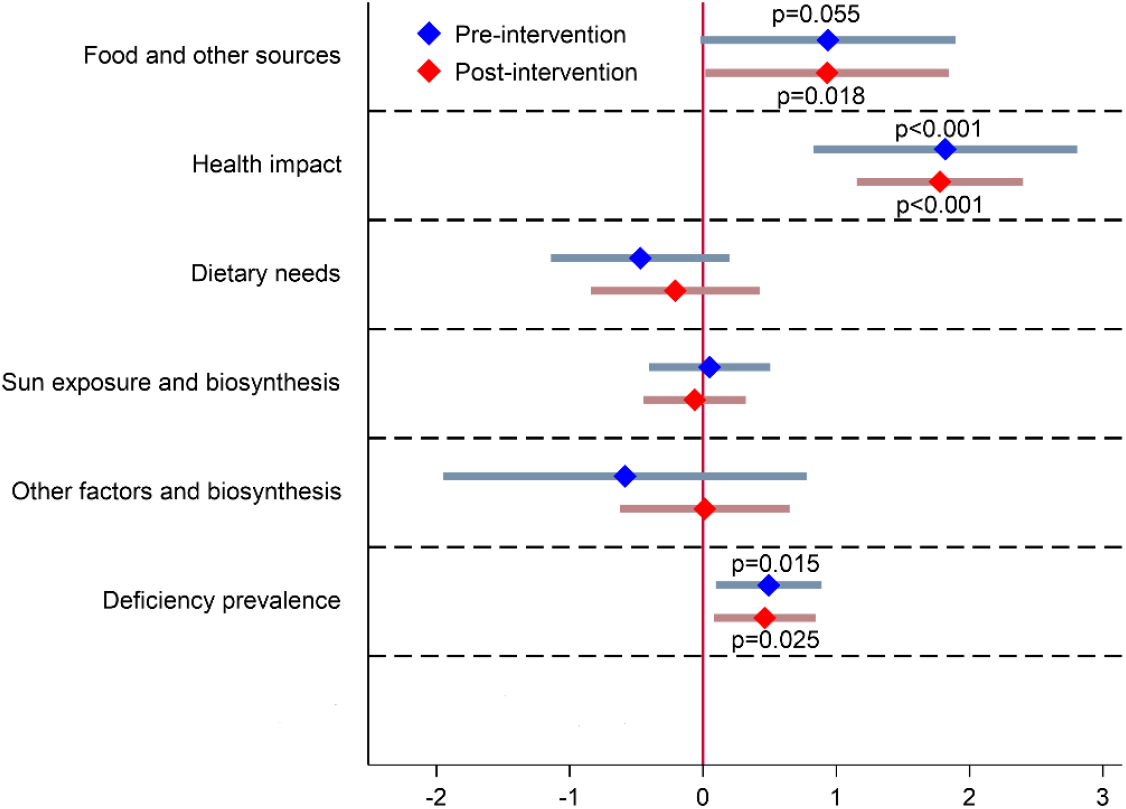
Logistic regression analysis for distinct dimensions of vitamin D-related knowledge in pre-intervention (Model 1: April 2020) and post-intervention (Model 2: December 2020) prevalence of vitamin D supplementation Note: Area under receiver operating characteristic (ROC) curve: 0.66 (pre-intervention) and 0.67 (post intervention)

## 4. Discussion

Due to previously established high prevalence of wintertime vitamin D deficiency in populations not taking vitamin D supplements (Calvo et al., 2005; Hribar et al., 2020; Pilz et al., 2018; Spiro & Buttriss, 2014), an educational intervention was conducted in Slovenia in November 2020, during COVID-19 pandemic. Objectives of this study were to investigate the voluntarily vitamin D supplementation practices, factors, that are affecting these practices, and to evaluate the effects of the educational intervention on vitamin D supplementation practices during the COVID-19 pandemic.

Interestingly, despite the very high prevalence of vitamin D deficiency in many countries, very few studies investigated the penetration of supplementation practices. While some countries introduced policies to implement vitamin D supplementation in specific and more at-risk groups (i.e., children up to 12 months of age in Slovenia), supplementation is typically voluntary in the general population. Spiro et al. (2014) highlighted major differences in the use of food supplements across Europe, with a clear north–south gradient. Typically, intake of food supplements is higher in northern countries. Greater use of food supplements is also commonly reported in women in comparison with men (Skeie et al., 2009). It has been established that, globally, dietary supplementation contributes 6–47% of the mean intake of vitamin D (Calvo et al., 2005). In a recent UK study, 43% of participants (adults) used vitamin D supplements (O’Connor et al., 2018); however, the study sample mostly included females and was not representative. Nevertheless, similar penetration of vitamin D supplementation was reported in Pakistani students (Tariq et al., 2020). On the other hand, a nationally representative French study reported the use of vitamin D supplements at a much lower level of 11% (Deschasaux et al., 2016). Similarly, only about 9% of adults reported year-round vitamin D supplementation in a Slovenian nationally representative dietary Si.Menu/Nutrihealth survey, conducted in 2017/2018 (Hribar et al., 2020). The same study also identified an alarmingly high prevalence of vitamin D deficiency (about 80%) in adults between the beginning of October and the end of April (extended wintertime), but the study design did not allow insights into the seasonal use of food supplements to be captured.

In April 2020, about one-third of our study sample reported extended wintertime vitamin D supplementation, and we did not observe considerable differences before and during the COVID-19 pandemic (33.7% vs. 33.2%, respectively). The observed greater penetration of vitamin D supplementation, in comparison with Si.Menu/Nutrihealth 2017/2018 data, can be partially explained by the fact that our measurements were done during the extended winter period when vitamin D supplementation is usually advised. Although at that time there were no official policy recommendations for vitamin D supplementation in the general population in Slovenia, this topic was addressed by the mass media in March 2020 (Figure 1), and greater penetration of the supplementation was expected during COVID-19 lockdown in April 2020. At that time some mass media reports were published (Siuka, 2020) about the importance of this vitamin for the functioning of the immune system, and about the possible beneficial role of vitamin D during the COVID-19 pandemic. However, there were no notable changes in the prevalence of vitamin D supplementation, or the daily dosages of vitamin D.

A very different situation was observed during the second wave of the COVID-19 pandemic, which affected Slovenia much harder. It should be noted that at the launch of the April 2020 survey, there were cumulatively 1,366 COVID-19 cases and 79 deaths reported in Slovenia, while in December there were already had 95,479 COVID-19 cases and 2,041 deaths (SIGOV, 2021). This also affected our study. In April 2020 survey, about 9% of participants reported that their household was somehow affected by COVID-19 (i.e. due to illness or quarantine of household member), while in December 2020 this was the case in 25% of subjects (Table 1). Furthermore, April 2020 mean score for the reported likelihood of a household member becoming infected with the virus was notably lower (score 2.2/5) in comparison with December 2020 measurement (score 2.7/5). Also, the December 2020 survey was conducted after the educational intervention. A press release (NUTRIS, 2020) about the wide prevalence of vitamin D deficiency in the Slovenian population (Hribar et al., 2020) was sent to all major mass media at the beginning of November 2020, which received a lot of media attention (Figure 1).

Analyses of the December 2020 dataset showed that penetration of dietary supplementation with vitamin D increased to 55.6% (from 33.7% in April), with the majority of supplement users taking a daily dosage of at least 25 µg vitamin D. The proportion of subjects with very high vitamin D intakes (above UL level of 100 µg/day (EFSA, 2012) increased during the pandemic, however about 95% of those supplementing vitamin D were still within safe intake levels (<100 µg/day). Nevertheless, the observation that, in some subjects, vitamin D intakes increased drastically during the pandemic highlights the need for very careful communication of vitamin D supplementation practices in relation to specific health-related events, such as the COVID-19 pandemic. It should be also noted that we recently investigated most commonly consumed Vitamin D supplements in the population (Žmitek, KruŠič, & Pravst, 2021). We analyzed 24 food supplements, which were purchased on the regular market on the market. Median labelled vitamin D (cholecalciferol) content was 25 µg and results of laboratory analyses confirmed expected amount of vitamin in majority (92%) of samples.

Vitamin D supplementation in the general population is likely to stay voluntarily in most countries. To use dietary supplementation as a strategy for lowering the risk of vitamin deficiency in such circumstances, very efficient public awareness programs would need to be implemented. In this study, we therefore focused on the identification of predictors of vitamin D supplementation practices. In April 2020 the most important predictors of vitamin D supplementation were the financial and health status of the participants, and specific dimensions of individual vitamin D-related knowledge. Multivariable logistic regression analysis highlighted the highest odd ratios for vitamin D supplementation in participants with a lower health status and in those with a higher financial status. In April 2020, only 33% of subjects used vitamin D supplements, while in the low health status group this was the case in 52%, and in the below-average financial status group in 25%. This indicates that the lowest supplementation rates were observed in those, who would probably need the supplementation the most. These observations are in line with our expectations that wealthier persons can more easily afford to purchase food supplements and that those with a lower self-reported health status more commonly used supplements. Interestingly, we did not observe significant differences between different sexes and age groups, although we would expect a higher penetration of supplementation in older adults (where vitamin D deficiency is commonly more pronounced (Spiro & Buttriss, 2014)), and in women, who are typically more frequent users of food supplements (Tariq et al., 2020). But the situation changed after the educational intervention; in December 2020 dataset age became the only strongly significant (P=0.01) parameter, with the highest supplementation rates in elderly subjects (72%). It should be also noted, that 52% of participants in the lower financial status group reported vitamin D supplementation. Study results indicate that we managed to considerably increase vitamin D supplementation across different population groups, including in the most vulnerable groups, such as the elderly population.

Similar to the observations of O’Connor et al. (2018) in the UK and of Boland et al. (2015) in Canada, subjects with better vitamin D-related knowledge are more likely users of vitamin D food supplements. Looking into different dimensions of vitamin D-related knowledge, logistic regression analysis highlighted three dimensions as independent predictors of vitamin D supplementation.

‐ (Q1) Dietary vitamin D sources: only a few foods are natural sources of vitamin D, and therefore dietary intake of vitamin D is typically very low (Lanham-New & Wilson, 2016). It seems that those who know that their diet is typically very poor in vitamin D are more likely to use vitamin D supplements.
‐ (Q2) Vitamin D health impact: vitamin D is a pro-hormone and is essential for musculo-skeletal health, normal immune system, and numerous other body functions (Autier et al., 2014; Zittermann, 2003; Zittermann et al., 2016). People that are more aware of these health-related functions are more likely to supplement their diet with vitamin D.
‐ (Q6) Prevalence of vitamin D deficiency: there is a very high prevalence of vitamin D deficiency in the population, particularly during the winter season (Hribar et al., 2020). Those that were aware of this fact can more easily consider themselves as at risk for vitamin D deficiency and are more likely to use vitamin D supplements. We should also note the previously reported strong inconsistency between personal opinions about Vitamin D status and actual vitamin D status (Deschasaux et al. 2016).

Interestingly, some vitamin D knowledge dimensions were not significantly connected to supplementation practices. For instance, knowledge about the (Q3) recommended daily amount of vitamin D, about the (Q4) time needed in the sun to get enough vitamin D, and about (Q5) factors that affect the skin’s biosynthesis of vitamin D were not independent predictors of vitamin D supplementation. This was noted both in pre- and post-intervention surveys. The above-mentioned observations are very important for the preparation of key messages that need to be well communicated if we want to increase vitamin D supplementation in the general population.

Our study also provides interesting insights into the overall knowledge about vitamin D in the Slovenian population. Knowledge was scored using a tool developed by Boland et al. (Boland et al., 2015). The original questionnaire was tested on Canadian students; the results showed poor knowledge and highlighted the need for more efficient health promotion programs. The reported mean total score in the Canadian study was 29%, while in our case it was 27% (1.60/6) before the intervention, and 37% (2.21/6) after the educational intervention. Knowledge about factors affecting vitamin D levels were also comparably low both in the Canadian study and in our pre-intervention April 2020 study (23%), but in our case this score increased to 45% after the educational intervention. On the other hand, in April 2020 we observed notably lower scores for vitamin D health impact than in the Canadian study (26% vs 37%, respectively), but this factor also notably improved after the intervention (40%). Contrary, about a quarter of our participants (both in pre-and post-intervention survey) correctly identified the amount of time in the sun required to produce adequate vitamin D (only 14% in the Canadian study), while, in both studies, less than 10% identified the correct recommended vitamin D intakes. It should be noted that other studies also identified serious vitamin D-related knowledge gaps in various other populations. Deschasaux et al. (2016) investigated vitamin D-related knowledge in a very large study in France, highlighting several knowledge gaps related to vitamin D sources and (non-skeletal) health effects. Tariq et at. recently investigated the vitamin D knowledge in Pakistani students (2020). Only 9% of study subjects correctly identified dietary sources of vitamin D, while one third were aware of the bone health-related effects of vitamin D, and only 36% identified sunlight exposure as a factor influencing vitamin D production. Interestingly, they also observed that those with more knowledge about the health functions of vitamin D were more likely to use vitamin D supplements. We should note, however, that there were considerable differences in the tools used for measuring vitamin D-related knowledge in these studies.

The strength of this study is in the controlled sampling conducted in two short duration periods during very early (April 2020) and late stage (December 2020) of the COVID-19 pandemic. While the use of an online panel could be considered as a study limitation, we should mention that considering pandemic-related restrictions, the use of an online study was the only option in practice. Both data collections were done during national lockdowns when all schools and universities were closed, non-essential workplaces in the public sector were closed and the private sector was recommended to close or restrict the number of people working; personal movement was restricted to within one’s municipality and operation of the public transport was limited. While food stores were open, non-essential stores were mostly closed. There was governmental advice in place to stay at home and to limit contact with others, while gatherings in public places were limited. We should note that, for some people, these circumstances might have limited the access of participants to vitamin D supplements during the COVID-19 pandemic. The quota sampling approach enabled a fair balance between the genders, age groups, and urban and rural areas. However, the approach used is also subject to limitations. The requirements for computer/smartphone use and internet access denied the inclusion of participants of the lowest socio-economic status. On the other hand, Slovenia has a very good internet infrastructure, and most households use computers. According to data from the Slovenian Statistical Office, more than 80% of the Slovenian population (16-74 years) is using internet (STAT, 2019). Also, home-schooling was in place in Slovenia during COVID-19 lockdowns for all elementary/secondary schools and universities, with online lectures. Nevertheless, the sampling approach may partially explain the difficulties in achieving representativeness in the study samples in the terms of educational level.

Another limitation is related to the vitamin D-related knowledge survey used. To provide some international comparability, we used a tool that was previously tested on Canadian students (Boland et al., 2015), but has not been validated or used in other subject groups. Despite the above-mentioned limitations, the authors believe that the tool used provided reliable predictors of vitamin D supplementation practices. We should also note that some of the study subjects participated both in April and December 2020 data collections. While this strengthens our study, because enabled us to investigate changes in the same subjects, such sampling could also present a limitation. Although study surveys were not conducted in a way to increase vitamin-D related knowledge or to affect vitamin D supplementation practices in study participants, survey questions brought vitamin D topic under attention. A series of control checks were therefore performed to verify, if this had any meaningful effect on the reported study results. For example, we have compared mean vitamin-D knowledge scores in the second survey (December 2020) between new subjects (N=233), and those that already participated in April 2020 survey (N=373), but no significant differences were observed. Furthermore, we compared December 2020 vitamin D supplementation prevalence in both these two groups. No meaningful differences were observed; in both groups vitamin D supplementation rates were above 50%, and median daily vitamin D intake was 25 µ. Therefore, we believe that the reported results were not majorly affected by exposing subjects to vitamin D topics.

## 5. Conclusions, policy, and research recommendations

While most foods are generally quite poor in vitamin D, they can assure adequate intake of this vitamin, particularly in regions with efficient food fortification policies (Calvo et al., 2005). However, in regions without such policies, a considerable proportion of the population is at risk for insufficient vitamin D status, which could be managed with supplementation. Findings of our study suggest that at beginning of 2020 most of the Slovenian population did not supplement their diet with vitamin D, despite the fact that previous studies indicated alarmingly high vitamin D deficiency prevalence between October and April. While we did not observe notable changes in vitamin D supplementation practices early in the COVID-19 pandemic (April 2020), in comparison to pre-COVID-19 observations, a very successful educational campaign using mass media resulted in a major increase in frequency of winter-time vitamin D supplementation. Pre-intervention study highlighted financial status as an independent predictor of vitamin D supplementation, with those with a below average financial status having the lowest proportion of vitamin D supplementation. This indicates that the financial dimensions of vitamin D supplementation also need to be considered by policy makers to ensure the protection of vulnerable groups.

Vitamin D-related knowledge was also found to be a key predictor of dietary supplementation, with some knowledge dimensions being more important than others. The three key dimensions identified as predictors of more likely vitamin D supplementation are knowledge about dietary sources of vitamin D, the health-related impact of vitamin D, and the prevalence of vitamin D deficiency in the population. Considering the study findings, the following key messages would need to be embedded into awareness campaigns in order to increase supplementation with vitamin D:

a. Vitamin D can be biosynthesized by human skin when we are sufficiently exposed to sunlight, but such biosynthesis is efficient only between May and September. (Note: this is geolocation-related information reflecting the situation in Slovenia).
b. In the absence of efficient biosynthesis, enough vitamin D needs to be provided by the diet. However, only oily fish and a few other foods are notable natural dietary sources of vitamin D. Therefore, the typical dietary intake of vitamin D with natural foods is much lower than recommended intake for the normal functioning of the human body.
c. Vitamin D has numerous health functions. It also contributes to the maintenance of normal bones, muscle function, and the function of the immune system.
d. In particular, between October and April, there is a very high prevalence of vitamin D deficiency in the population. (Note: this is nationally specific information reflecting the situation in Slovenia).

These key messages were constructed based on our preliminary results, using April 2020 sampling, and used in the populational educational intervention in November 2020. Herein reported study results showed that the intervention was very efficient, however long-term effects are yet to be determined in future seasons. Additional studies are therefore needed in the future, preferably in a similar calendar season – during the winter. If vitamin D supplementation practices will change in long term, epidemiological data on vitamin D status in key population groups should be also revisited. It should be noted that the above provided communication messages result from data collected in the Slovenian population. While very similar messages might also be applicable in other regions, they should be adapted to address regional and population differences. The efficiency of awareness campaigns should be always carefully evaluated.

## Data Availability

The raw data supporting the conclusions of this article will be made available by the authors, without undue reservation.

## Ethics Statement

Ethical approval for this study was obtained from the Bioethical Committee of the Higher School of Applied Sciences in Ljubljana, Slovenia (VIST ET-6/2020). The survey was conducted in the Slovenian language as an amendment of the international survey Food-COVID-19: Lifestyle and dietary patterns, and nutrition knowledge. The participants provided their informed consent to participate in this study via an online form.

## Author Contributions

I.P. and K.Z. conceived the study. I.P., K.Z., M.H. and A.K. designed the study questionnaire. K.Z., H.H., and I,P. analyzed and interpreted the data. M.H. and Z.L. supported with the data analyses, and H.H. conducted the statistical analyses. K.Z. wrote the first manuscript draft and all authors then made revisions. All authors have read and approved the final version of the manuscript.

## Acknowledgments

We acknowledge the support of Nina Zupanič and Sanja Krušič (Nutrition Institute, Ljubljana, Slovenia) in the testing of the survey tool, and the support of Mr. Primož Logar and the team at Marketagent reSEARCH GmbH (Baden, Austria) for the programming of the survey tool and the use of their consumer panel in Slovenia. We also acknowledge the support of physicians in communications with media and practitioners, particularly Marija Pfeifer, Darko Siuka, Barbara Hrovatin, Bojana Pinter, Zvonka Slavec and Alojz Ihan.

